# Personalised prediction of daily eczema severity scores using a mechanistic machine learning model

**DOI:** 10.1101/2020.01.16.20017772

**Authors:** Guillem Hurault, Elisa Domínguez-Hüttinger, Sinéad M. Langan, Hywel C. Williams, Reiko J. Tanaka

## Abstract

**Background:** Atopic dermatitis (AD) is a chronic inflammatory skin disease with periods of flares and remission. Designing personalised treatment strategies for AD is challenging, given the apparent unpredictability and large variation in AD symptoms and treatment responses within and across individuals. Better prediction of AD severity over time for individual patients could help to select optimum timing and type of treatment for improving disease control.

**Objective:** We aimed to develop a mechanistic machine learning model that predicts the patient-specific evolution of AD severity scores on a daily basis.

**Methods:** We designed a probabilistic predictive model and trained it using Bayesian inference with the longitudinal data from two published clinical studies. The data consisted of daily recordings of AD severity scores and treatments used by 59 and 334 AD children over 6 months and 16 weeks, respectively. Internal and external validation of the predictive model was conducted in a forward-chaining setting.

**Results:** Our model was able to predict future severity scores at the individual level and improved chance-level forecast by 60%. Heterogeneous patterns in severity trajectories were captured with patient-specific parameters such as the short-term persistence of AD severity and responsiveness to topical steroids, calcineurin inhibitors and step-up treatment.

**Conclusion:** Our proof of principle model successfully predicted the daily evolution of AD severity scores at an individual level, and could inform the design of personalised treatment strategies that can be tested in future studies.

## INTRODUCTION

Atopic dermatitis (synonymous with atopic eczema or just eczema [1]; AD) is the most common inflammatory skin disease, and is characterised by inflamed, dry and itchy skin [2] leading to substantial quality of life impairment and significant socioeconomic impact [3]. AD typically has a fluctuating course characterised by inflammatory disease flares followed by periods of remission. Treatment with topical corticosteroids or calcineurin inhibitors during disease flares is aimed at controlling symptoms and skin signs, and emollients are typically used to counteract the dry skin associated with AD.

However, successful control of AD symptoms has been challenging as responses to AD treatments vary considerably between patients. Personalised treatment strategies may be more beneficial to individual patients rather than a “one-size-fits-all” approach to therapy [4] [5]. A first step toward developing personalised treatment strategies is to better predict the consequences of possible treatments at an individual level, rather than at population level, to deal with the variability across patients.

Prediction of the consequences of treatments at an individual level is challenging also because of dynamic and sudden fluctuations of AD symptoms. It can be difficult to identify reliable treatment responses, especially if a single endpoint is considered, since the responses to a treatment can vary each time even for the same patient. Analysing the dynamic responses to the repeated application of treatment can help identify consistent treatment effects for each patient [6] and ultimately predict whether the chosen treatment is effective and whether the disease is adequately controlled at an individual level.

Machine learning has been successfully applied for prediction tasks. However, typical machine learning models such as artificial neural networks are often black-boxes, lacking interpretability, or relying on *post-hoc* explanations that are not guaranteed to match the algorithm’s true decision process [7] [8]. “Black-box models” may fail to be accepted by the medical community and AD patients. Existing regulations such as the European Union general data protection regulation also highlights the “pressing importance of human interpretability in algorithm design” [9].

Here we aimed to develop a biologically interpretable mechanistic machine learning model that can predict daily evolution of AD severity scores at an individual level. We applied a model-based machine learning approach [10], which allowed us to develop a high performing Bayesian machine learning model that can be tailored to the particular context of a given study and the available dataset, and include biologically interpretable mechanistic knowledge. Bayesian machine learning approach has already been applied to a birth cohort data on allergic sensitisation to uncover latent atopy classes [11] or to estimate asthma misclassification and risk factors in yearly questionnaire data [12]. However, it has not been applied to predict *daily* changes in disease outcome or in the field of AD.

We hypothesised that it is possible to decipher the apparent unpredictable dynamics of AD severity scores from each patient’s data. We previously published a mechanistic model of AD pathogenesis which provided a coherent mechanistic explanation of the dynamic onset, progression, and prevention of AD, as a result of interactions between skin barrier, immune responses and environmental stressors [13] [14]. Our aim was therefore to adapt the structure of the published mechanistic model to real patient data (Fig S1), and to develop a mechanistic Bayesian model tailored to each individual, that can predict the next day’s AD severity score given their score and treatments used on that day.

## METHODS

### General approach

Using the longitudinal data from two published clinical studies [15] [16] (Fig S1), we developed and validated a mechanistic Bayesian model that can predict the next day’s AD severity score for each patient. Our mechanistic Bayesian model explicitly described within-patients uncertainties in disease outcomes using probability distributions, and between-patients heterogeneity in severity trajectory and treatment responses by patient-dependent parameters.

To develop the model, we firstly defined the underlying processes that could generate the data as a probabilistic graphical model (Fig 1A), which adopted the structure of a previously published mechanistic model of AD pathogenesis [13] [14] (Fig 1B). The model was tailored to the context of the clinical studies in which the data was collected. We then trained the model (fitted to the data) using Bayesian inference, i.e. updating the probability distributions of the unknown variables and model parameters through Bayes’ theorem, and validated the model by assessing its predictive performance in a forward chaining setting, where the model was trained with the first week’s data and tested on the second week’s data, then re-trained on the first two weeks’ data and tested on the third week’s data, and so on (Fig S2). The first dataset was used for model development and internal validation, and the second dataset for external validation to ensure its generalisability to a similar but independent population.

**Figure 1:**
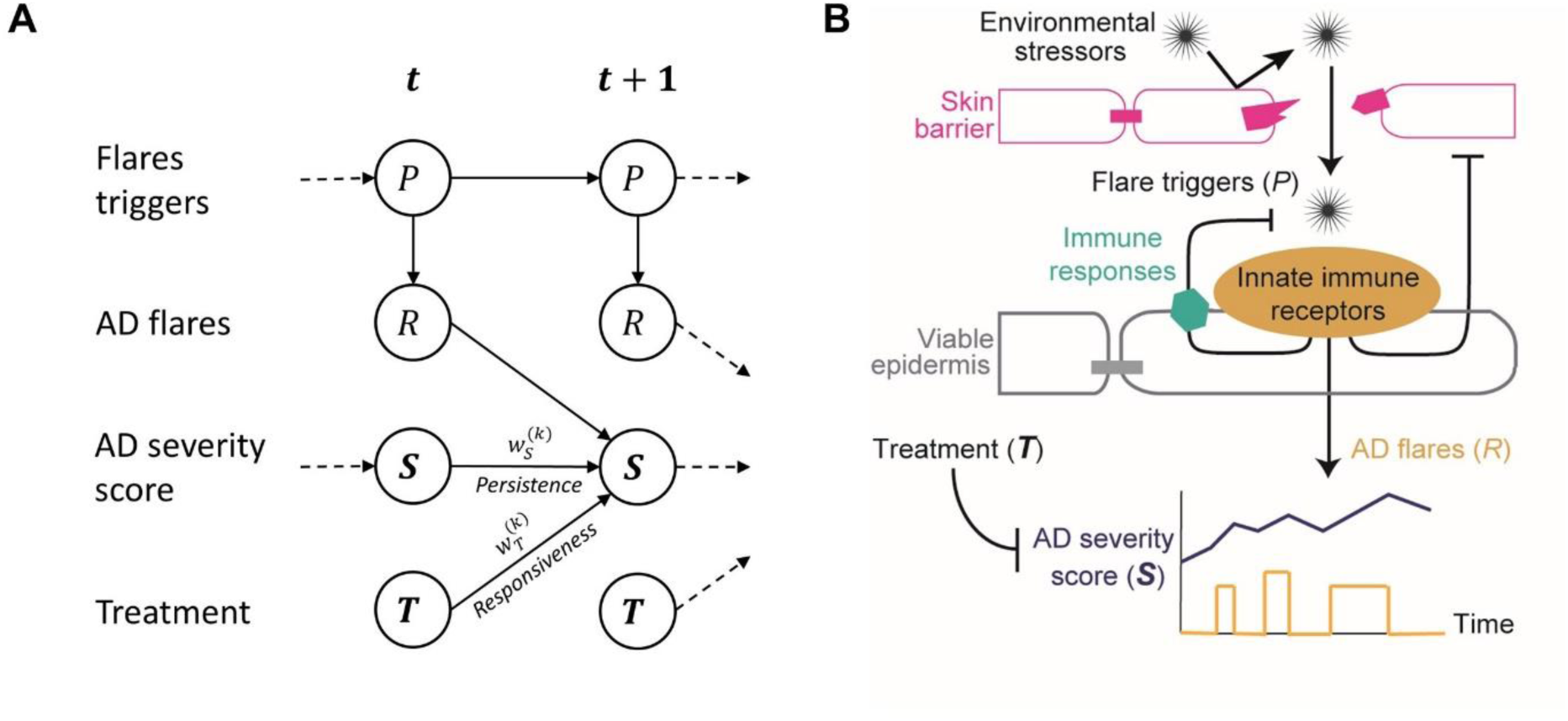
Mechanistic Bayesian model of AD severity dynamics. A: Probabilistic graphical model representation. The arrows depict the relationships between state variables included in the model. B: A schematic diagram of the published mechanistic model of AD pathogenesis [13] from which the structure of the proposed model was adopted.

### Data

We chose two datasets that included daily recording of symptoms and treatments over a moderately long period.

The first dataset, which we refer to as “Flares dataset”, is a part of the data collected in an observational study that aimed to identify the triggers of AD flares for 59 children [15]. The Flares dataset included *daily* categorical “bother” scores over 6 to 9 months, totalling 6536 patient-day observations, graded from 0 (“no bother at all”) to 10 (“the most bother you can imagine”) as a response to the question “how much bother did your eczema cause today?”. 38.8% of the bother score was missing in Flares dataset (Fig S3). The Flares dataset also included daily binary “stepping up” variables, i.e. the answers to the question “have you had to step up your treatment today because your eczema was worse?”. What constituted “stepping up” treatment was defined for each child at the study outset.

The second dataset, which we refer to as “SWET dataset”, is a part of the data collected in a randomised controlled trial that evaluated the effects of use of ion exchange water softeners for AD control (the softened water eczema trial or SWET) for 334 children [16]. The SWET dataset included the individual child’s daily categorical bother score over 16 weeks with only 1.9% of the bother score missing (Fig S4) for a total of 35854 patient-day observations. The SWET dataset additionally contained information on potential risk factors or confounders, such as the presence of filaggrin mutations, white skin type, age (in years), gender, and whether the patient slept away from home. It also included details of the treatment used, such as the type of treatment modalities used each day (topical corticosteroids, calcineurin inhibitors and stepping-up treatment), the estimated average dose used for each type of topical corticosteroids (mild, moderate, potent or very potent) and calcineurin inhibitors (mild or moderate) over the study period, together with the patient’s confidence in the estimated average dose (“not at all sure”, “not sure”, “sure”, or “very sure”). We used all the available information in SWET dataset and evaluated the contribution of each factor on daily evolution of the bother score at an individual level.

### Mechanistic Bayesian models

We developed a mechanistic Bayesian model that predicts the AD severity score (***S***_*k*_(*t* + 1)) for the *k*-th patient at day *t* + 1, given the previous day’s score (***S***_*k*_(*t*)) and the treatment applied (***T***_*k*_(*t*)) (Fig 1A). Our model assumed that the AD severity score is continuous, determined by the temporal accumulation of inflammation caused by AD flares (*R*_*k*_(*t*)) and modified by the treatment applied (***T***_*k*_(*t*)), and that AD flares result from activation of innate immune receptors by flare triggers (*P*_*k*_(*t*)) (Fig 1B). Flare triggers (*P*_*k*_(*t*)) and the resulting flares (*R*_*k*_(*t*)) depend on the complex interactions between the skin barrier, immune responses and environmental stressors, and were modelled as latent variables. The recorded and non-recorded variables are shown here in bold and non-bold, respectively.

We described the dynamic evolution of the AD severity score by an exponentially modified Gaussian distribution, 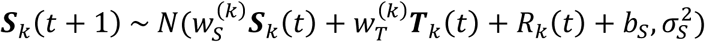, that is, the distribution of ***S***_*k*_(*t* + 1) follows a Gaussian autoregressive process perturbed by exponentially distributed AD flares, *R*_*k*_(*t*) ∼ *Exp*(*β* = *P*_*k*_(*t*)), which reflects the assumption that flares occur more frequently in the presence of the flare triggers. The autoregression is characterised by the patient-dependent autocorrelation or persistence of the severity score 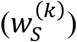 and population-level intercept (*b*_*s*_) and variance 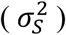. The model is adjusted for the treatment applied with the patient-dependent responsiveness to treatment 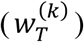 sampled from the hierarchical prior 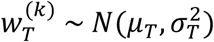 with population mean *μ*_*T*_ and variance 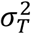. We modelled the relative change of the amount of flare triggers (*P*_*k*_(*t*)) by a Gaussian random walk, 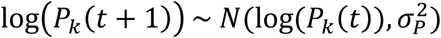, of variance 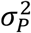.

We also developed an extended version of the mechanistic Bayesian model for SWET dataset (details in Supplementary A). The extended model allowed us to analyse the effects of potential risk factors (the presence of filaggrin mutations, age, and sleeping away from home) on the severity score, with their respective weighting parameters, 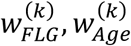, and 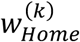. We also investigated heterogeneity of treatment responsiveness by replacing the term 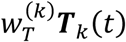 with 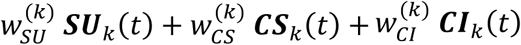, where ***SU***_*k*_ (*t*), ***CS***_*k*_ (*t*) and ***CI***_*k*_ (*t*) are binary variables that indicate whether the *k*-th patient stepped-up, applied topical corticosteroids and calcineurin inhibitors, respectively, with their respective weights, 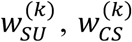 and 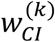. The weights, 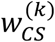 and 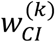, include dose-independent effects (intrinsic responsiveness to the treatment 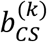 and 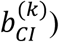 and dose-dependent effects that are functions of the quantity and the potency of the treatment (Fig S5).

Missing values in ***S***_*k*_(*t*) were treated as parameters to be inferred by the model in a semi-supervised setting (detailed in Supplementary B). The choice of priors and hierarchical priors are described in Supplementary C.

### Model fitting

Model training was performed using the Hamiltonian Monte-Carlo algorithm in the probabilistic programming language Stan [17]. The posterior distribution was sampled by 6 Markov chains for 3000 iterations (including 50% burn-in). Convergence of the chains was monitored by inspecting the trace plots, checking the Gelman-Rubin convergence diagnostic 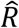 [18] and computing effective sample sizes. More details of the inference method are provided in Supplementary D.

### Model validation

Internal validation was performed on Flares dataset to assess predictive performance on observations with which the model was developed. The SWET dataset was used to test the external validity (transportability) of the learning algorithm [19], i.e. whether the model can be updated from a set of priors and achieve similar performance to the development dataset (rather than testing the performance of a frozen model).

The predictive performance of the model was assessed in a forward chaining setting, in which we applied multi-category calibration using pairwise one-against-all isotonic regression (detailed in Supplementary E). Model calibration (whether forecast probabilities are accurate) was assessed by a normalised quadratic scoring rule (ranked probability skill score, RPSS) and discrimination (whether the ranking of probabilities is accurate) by the area under the receiving operating characteristic (AUROC). These metrics were plotted against training day (training data size) to produce learning curves. Details on performance metrics used are described in Supplementary F.

## RESULTS

### Model fitting

The model was trained on each of the two datasets and we checked the convergence. Population-level parameters (parameters shared across patients) were estimated with a good precision and their 95% credible interval (in which the parameter lies with 95% probability) were narrow compared to the mean, did not include 0 and were similar for the two datasets, suggesting support for the model structure (Table S1). Two main model parameters that describe patient-dependent dynamics of the severity score are the autocorrelation parameter 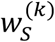 for the short-term persistence of the AD severity score, and the parameter 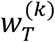 for the responsiveness to treatment, of the *k*-th patient. These estimates greatly varied from one patient to another, confirming their patient-dependence (Figs S6 and S7).

Posterior predictive checks for the severity score dynamics demonstrated that the developed model captured diverse patterns of the trajectories despite the presence of missing values (representative patients’ score dynamics in Fig 2, top). Typical trajectories observed included fluctuations of the severity score with a return to a healthy state (Fig 2A) or without (Fig 2B). Trajectories of the latent variable for the flare triggers (*P*_*k*_(*t*)) indicated a long-term dynamic trend of the severity score, with a decrease in *P*_*k*_(*t*) corresponding to less frequent flares (Fig 2, bottom).

**Figure 2:**
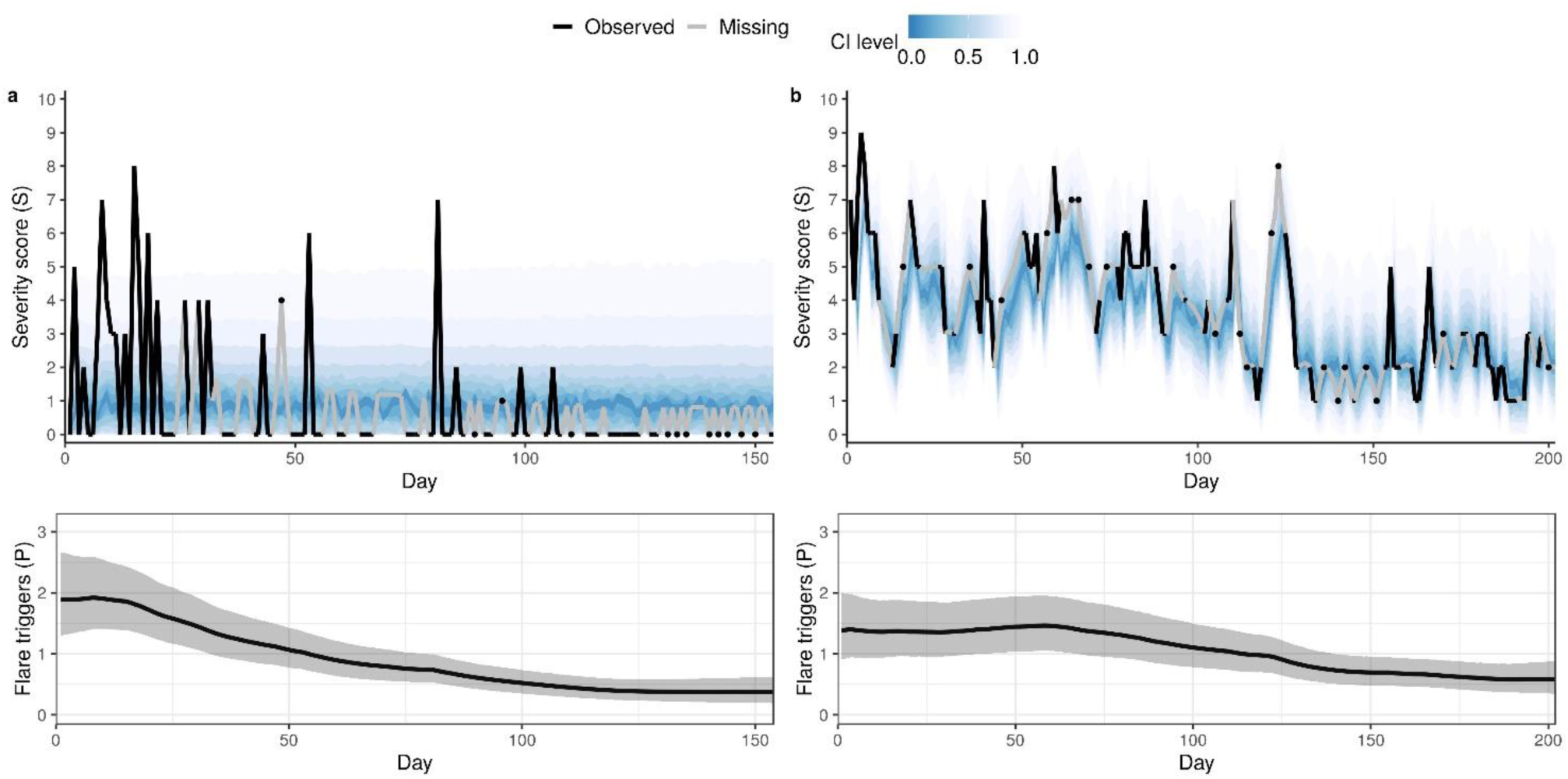
Fitting of the mechanistic Bayesian model. Posterior predictive distribution of AD severity score (top) and the corresponding trajectory of the latent variable for flare triggers (bottom) for two representative patients from Flares dataset. (A) The bother score returns to a healthy state with less frequent flares, accompanied by a decrease in flare triggers. (B) The bother score does not return to a healthy state despite the decrease in flare triggers. The top panel shows a fan chart of the posterior predictive distribution, where the highest density intervals corresponding to different credible levels are overlaid. The intervals are centred around the maximum a posteriori (mode), with darker colour corresponding to narrower intervals. Black and grey lines show the observed scores and the posterior mean estimate for the missing scores, respectively. In the bottom panel, the black line and shaded area represent the posterior mean of *P*(*t*) and 90% credible interval, respectively.

### Model validation

We then validated the model to assess its generalisability beyond the training data. The learning curves demonstrated an improvement in both calibration and discrimination, as more data becomes available (Figs 3A-B), confirming that the model learned the dynamic patterns of the severity scores from the data. The reliability of our model predictions was improved by adjusting the forecasts with multi-category calibration (Fig S8). Internal and external validity was confirmed by a calibration 48% and 61% better than a chance-level forecast and a discrimination of 87% and 90% on Flares and SWET datasets, respectively. Predicted probabilities up to 30-40% in Flares dataset and up to 50-60% in SWET dataset match observed frequencies (Figs 3C-D), with prediction for low bother scores being slightly underconfident in SWET dataset. We also implemented a Gaussian random walk as a null model and showed that our model outperformed it overall (details in Supplementary G).

**Figure 3:**
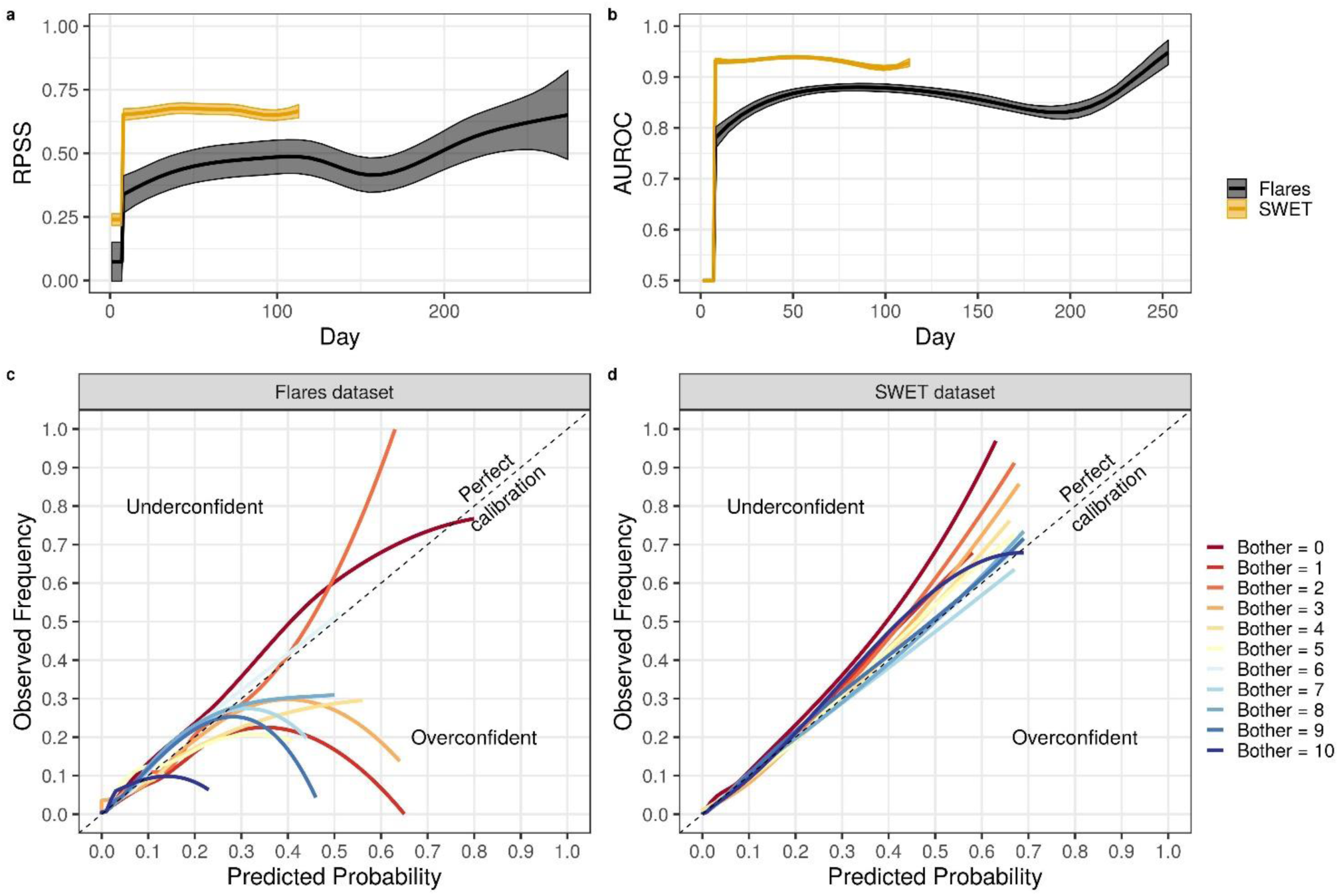
Predictive performance evaluation of the mechanistic Bayesian model. A-B: Evolution of calibration measured by RPSS (A) and of discrimination measured by AUROC (B) for one-day-ahead predictions as a function of the training day for Flares (black) and SWET (orange) dataset. As time (Day) increases, more data becomes available for training. A score of 1 corresponds to a perfect calibration/discrimination and 0 to a calibration/discrimination not better than chance. C-D: Calibration curves for the models trained by Flares dataset (C) and SWET dataset (D), obtained using locally weighted scatterplot smoother (LOWESS).

Similar results were obtained for a model we developed using the daily scratch score recorded in the observational study for Flares dataset (Fig S9). The scratch score was not recorded in SWET.

### Effects of treatment modalities and risk factors on the predicted severity scores

The extended model with additional covariates was also successfully fit to SWET dataset (Tables S1 and S2). The posterior predictive checks confirmed that the model could capture diverse patterns of the severity score trajectories, such as large and rapid fluctuations (Fig 4A), large but slow fluctuations (Fig 4B), and controlled AD (Fig 4C). The model could not predict previously unseen patterns, such as transitions of the score from 1 to 10 in a day (Fig 4D at around 70 days), as the model learned the dynamic patterns from past data.

**Figure 4:**
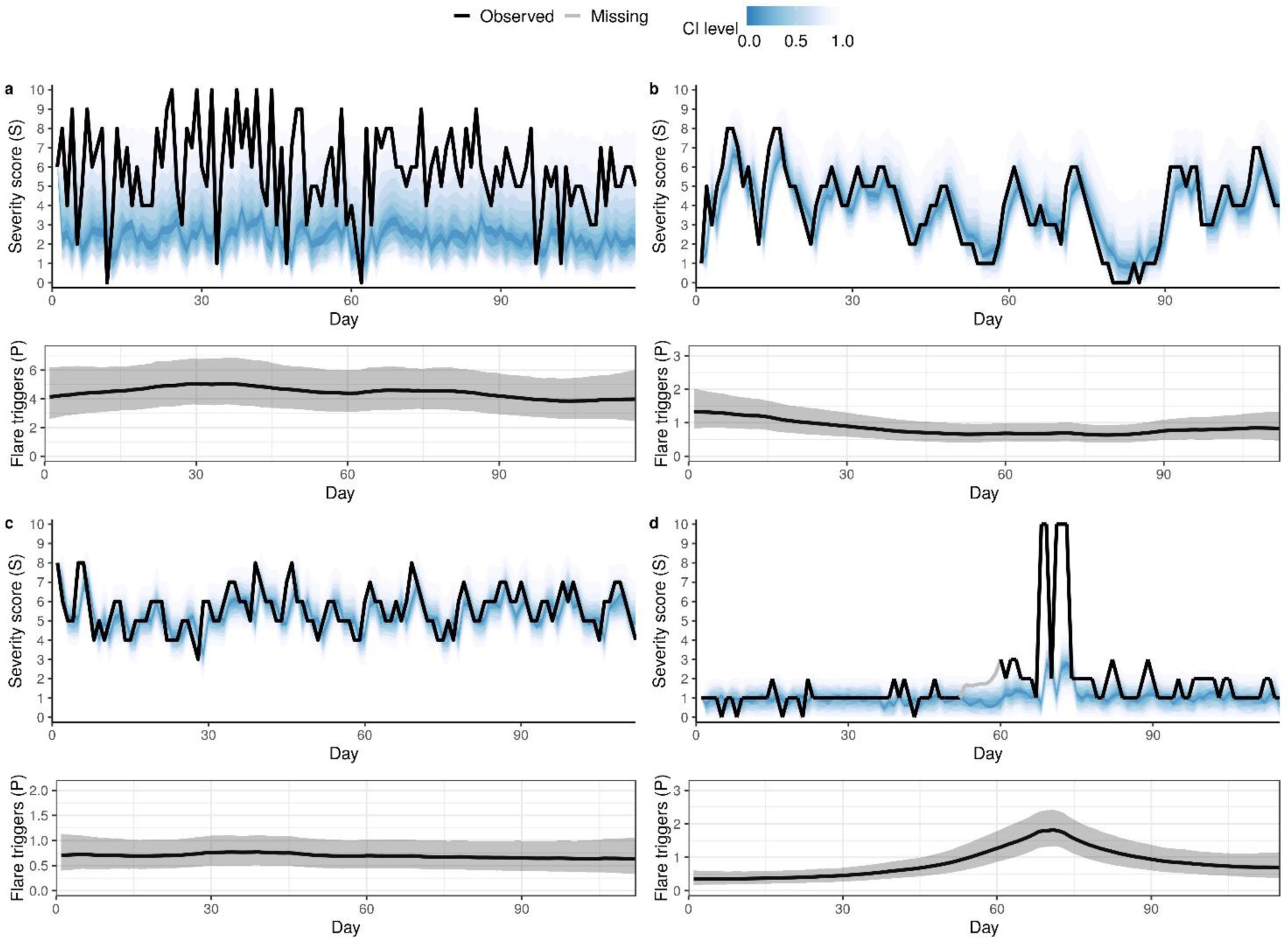
Fitting of the extended model. Posterior predictive distribution of AD severity score (top) and the corresponding trajectory of the latent variable for flare triggers (bottom) for four representative patients from SWET dataset: (A) large and rapid fluctuations, (B) large but slow fluctuations, (C) controlled AD, and (D) controlled but with transitions of the score from 1 to 10 in a day (at around 70 days). The top panel shows a fan chart of the posterior predictive distribution, where the highest density intervals corresponding to different credible levels are overlaid. The intervals are centred around the maximum a posteriori (mode), with darker colour corresponding to narrower intervals. Black and grey lines show the observed scores and the posterior mean estimate for the missing scores, respectively. In the bottom panel, the black line and shaded area represent the posterior mean of P and 90% credible interval, respectively.

Analysis of the model parameters suggested that older age, absence of filaggrin gene mutations, and sleeping at home were associated with *greater* improvement (decrease) in severity scores at the 95% credible level (Fig 5A), as the 95% credible interval of the relevant parameters (*w*_Age_ < 0, *w*_FLG_ > 0, and *w*_Home_ < 0) did not contain 0. The estimated effects may appear small (in absolute terms), compared to the range of the bother score (0-10), but their effects on the severity score may become practically significant as they accumulate over time. White skin type and sex were not found to be associated with changes in the severity score at the 95% credible level (Fig 5A; 95% credible interval of *w*_Sex_ and *w*_White_ in both sides of 0).

**Figure 5:**
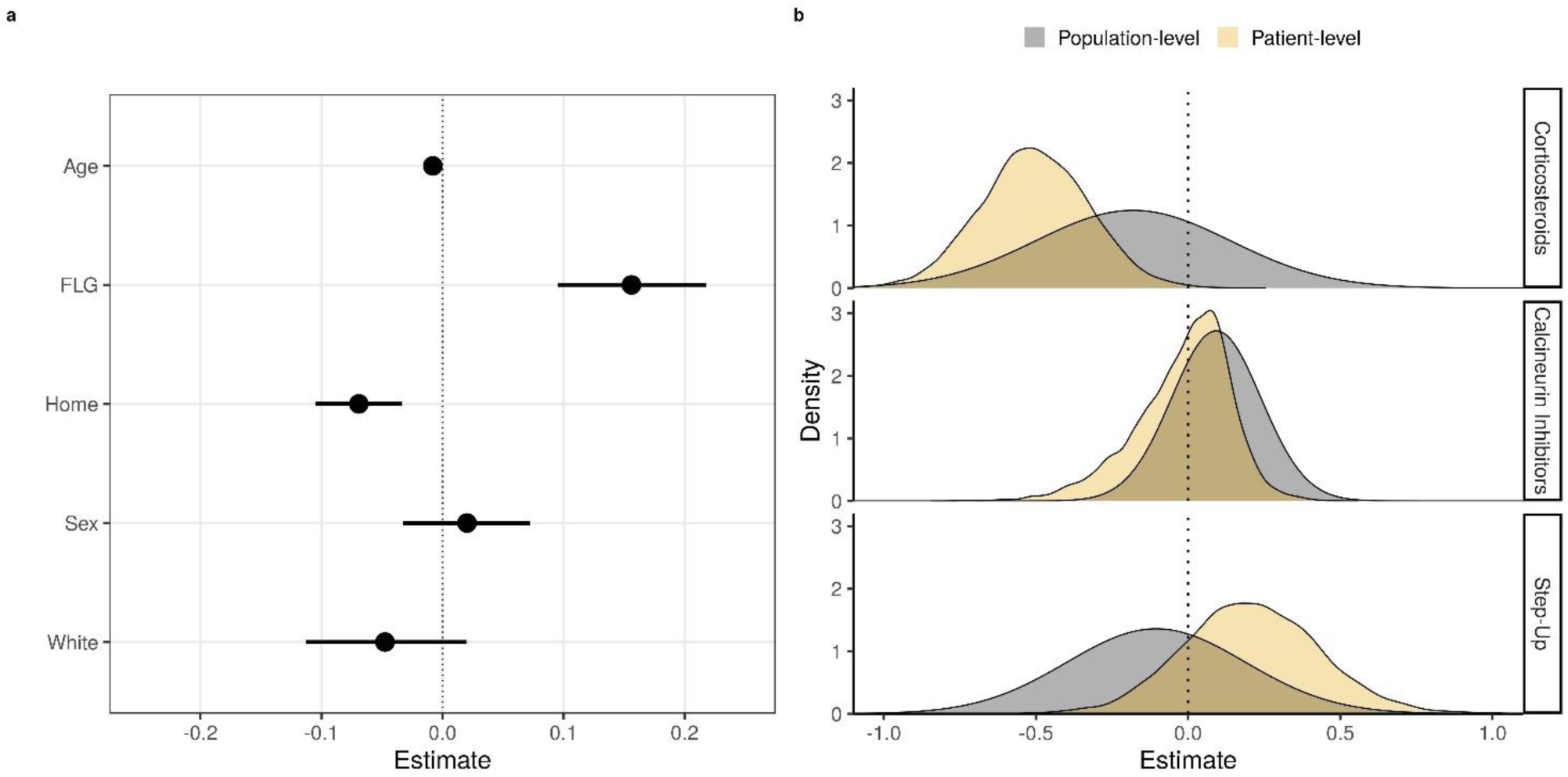
Estimated effects of potential risk factors and responsiveness to treatments on the severity score. A: Population-level estimates of the parameters 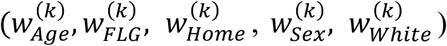 for potential risk factors (age, presence of filaggrin mutation, sleeping at home, sex, and white skin). The values represent the contribution of the relevant factor to the severity score. Negative and positive values represent a decrease and an increase in severity score (improvement and worsening), respectively, while null values suggest an absence of an effect. Black circles and the line segments represent the mean posterior and the 95% credible interval, respectively. B: Estimated distribution of the parameters for dose-independent responsiveness to different treatment modalities (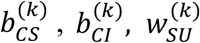 for corticosteroid, calcineurin inhibitors and step-up) at a population-level (grey) and for a specific patient (orange).

Further analysis of the parameters, 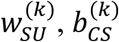 and 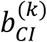, which describe the dose-independent effects of the treatment on the severity score, demonstrated that none of the treatments appears to have a significant effect at the population level (grey shaded areas in Fig 5B spans from negative to positive values). However, the treatments could have a significant effect at a patient-level. For example, the parameter estimates for one of the patients (orange shaded areas in Fig 5B) suggest that the use of corticosteroids has a significant and consistent effect on the severity score for this patient at the 95% credible level. That is, the posterior probability for 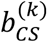 (the dose-independent responsiveness to corticosteroids) being negative (i.e. the use of corticosteroids reduces the severity score) is greater than 95%. Interestingly, this 95% criterion for the consistent treatment effect was not met for calcineurin inhibitors 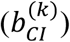 and step-up 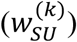 for the same patient. Following this criterion, we confirmed significant effect of corticosteroids in 70 individuals (out of 295 who used corticosteroids) and of step-up in 27 individuals (out of 284 who used step-up). However, we did not find evidence of an intrinsic responsiveness in any of the 92 patients who used calcineurin inhibitors, although 4 of them show a significant dose-dependent responsiveness.

## DISCUSSION

### Main findings

This study demonstrated a proof-of-concept that predicting the evolution of eczema severity is possible. We developed a novel mechanistic Bayesian machine learning model that can predict patient-specific daily evolution of the AD bother score. The model is biologically interpretable and describes the mechanistic assumption that the AD severity is a result of temporal accumulation of flares (Fig 1). The model learned rich, heterogeneous and dynamic patterns in the daily evolution of AD severity scores, that may otherwise appear random and noisy (Figs 2 and 4, top). Our method extracted information on whether the chosen treatment is effective (responsiveness to treatment) and whether the AD score is persistent, at an individual level (Figs 5, S6 and S7). The use of longitudinal data enabled us to look for consistent treatment responses within each patient, rather than a population average response valid at a single time point. We also inferred the long-term trend of the severity score dynamics (Figs 2 and 4, bottom), and estimated population-level risk factors associated with slower improvement of the severity score, such as the presence of a filaggrin mutation and younger age (Fig 5A). The model was validated using the data from two published clinical studies to confirm its generalisability and the possibility to learn and predict the short-term dynamics of AD severity scores from each patient’s data (Fig 3).

### Strengths of our approach

Our model-based Bayesian approach is useful to develop models for clinical use, especially when the data is not as controlled as in a clinical trial. Our model explicitly describes uncertainties in disease outcomes (the severity scores) using probability distributions rather than point estimates. The Bayesian approach also enabled us to deal with the missing data (about 40% of scores were missing in the Flares dataset) by treating them as parameters in a semi-supervised learning setting, where the model imputes the missing data while learning the dynamic patterns of the severity scores from the available data. This method is particularly appropriate for incomplete and partially missing data, for example, when patients miss clinical visits.

The model-based approach allows us to design models by taking prior clinical and mechanistic knowledge into account, and by tailoring them to available data and study context. For example, our model was extended according to the additional information (on potential risk factors and the treatment doses) available in SWET dataset but not in Flares dataset. When similar Bayesian models are developed, our parameter estimates can be incorporated as prior information, as the model can be learned in a sequential manner as new data comes in. For example, the model developed in this study could be expanded to include additional predictors or new patients, if relevant data becomes available, using the population posterior distribution of the patient-dependent parameters obtained here.

These features entail that the developed model cannot be made readily available as a “plug-in” formula, as it is described by a set of context-dependent equations on probability distributions and patient-specific parameters.

### Limitations of the study and future directions

The datasets we used in this study contained daily measurement of the bother score, a subjective global measure of distress caused by AD that has previously been used as a reference for developing asthma severity instruments [20] and validating AD symptom measures such as POEM [21]. While using objective and quantitative measurements would be preferable, this study can serve as a proof-of-concept that predicting the evolution of eczema severity is possible. When collecting daily measurements of objective severity scores becomes less challenging, similar models could be developed to predict scores such as EASI [22], (o)SCORAD [23], or their self-assessed versions. It will allow us to evaluate the dynamics of scores that capture different aspects of AD symptoms and to compare the predictive performance for different scores. It is also possible to investigate longer time horizon with weekly (instead of daily) measurements. Appropriate evaluation of the effects of data frequency on score dynamics prediction will help designing more effective and informative clinical trials towards personalised medicine.

The predictive capabilities of the model could be potentially improved with more data, by including additional predictors such as environmental or biological markers, or by using better-quality data, i.e. with fewer missing values or more precise information about treatments. For example, our model could not estimate with certainty the true quantity of treatment applied, and thus the relative effectiveness of different potencies of treatment, confidently from the available data (Table S2). We believe that the uncertainty was due to a potential difference between the reported average dose and the actual daily dose for each treatment and patient. The daily record of the quantity of treatment applied could resolve this issue and lead to a better estimate of treatment responsiveness.

The model proposed in this paper adopted a structure that was tailored to the available datasets. The model structure was much simpler than that of the previously published mechanistic model of AD pathogenesis [13] [14]. If the longitudinal measurement for interactions between environmental stressors, the skin barrier and immune responses becomes feasible in future, such data can be incorporated to develop a more detailed mechanistic machine learning model that provides deeper biological interpretation.

The model-based machine learning approach demonstrated here is applicable to help quantify patient responses to treatment, and may be suitable as a computational method for therapeutic stratification by identifying treatment responses for each individual [24]. The prediction of daily evolution of severity scores could be further used to suggest optimal treatment strategies for individual patients, using reinforcement learning for example, in addition to conventional computational methods using optimal control theory and bifurcation analysis [12] [25]. Our method could be tested further as part of an intervention using a personalised approach in a future pragmatic randomised controlled trial, and compared with conventional standard approaches.

## Data Availability

All the codes will be made available on GitHub.

## DATA AVAILABILITY

All the codes will become available on GitHub at the time of online publication.

## ACKNOWLEDGEMENTS

We thank Professor Kim S Thomas for sharing SWET dataset and constructive comments on the manuscript. The SWET trial was funded by the NIHR Health Technology Assessment Programme.

